# Identification of SARS-CoV-2 Persistent Intestinal Epithelial Syndrome (SPIES) as a Novel Disease Entity using Clinical, Histologic, and RNA Programmatic Data

**DOI:** 10.1101/2024.07.18.24310647

**Authors:** Thomas Wallach, Ahmed Soliman, John Agboola, Shagun Sharma, Lais Araujo-Coelho, Meredith Pittman, Christos Chatzinakos, Sergios-Orestis Kolokotronis

## Abstract

“Long COVID” (LC) remains an ongoing issue and one which has created a substantial burden of disease. Gastrointestinal LC is relatively poorly understood. In this study we characterize a syndrome of persistent SARS-CoV2 viral material via clinical and histologic data, and RNA sequencing

**Methods:** We reviewed patients aged 5-22 years with an esophagogastroduodenoscopy (EGD) for gastrointestinal (GI) symptoms from 6/2020-6/2023, excluding patients with known histologic disease. Biopsies were sent for immunohistochemical staining. Clinical data was collected. Duodenal, ileal, cecal, and sigmoid colon samples were stained for SARS-CoV-2 using a SARS-CoV-2 nucleocapsid antibody. Slides were reviewed by a blinded pathologist. 8 patients with known duodenal SARS-CoV-2 nucleocapsid antigen (SC-NA) positivity and 8 demographically matched IBS matched patients from prior to 2020 were identified for RNA sequencing comparison. Results were compared with public data from the Gene Expression Omnibus (GEO) data repository for intestinal tissue with IBS and epithelial tissues with active SARS-CoV2 infection.

**Results:** Of 30 patients, fifteen (50%) were identified to have positive SC-NA. 3 (20%) had received at least a single SARS-CoV2 vaccine in the + cohort, and 8 (53.3%) in the - (P=0.05). Primary symptoms were pain (86%, nausea (66.6%), and weight loss (60%). 37.5% of patients with colonic SC-NA displayed hematochezia. 33% of + patients showed elevated ESR/CRP. Mean + calprotectin was 317.3 vs. 156.4 (p=0.2). 11/15 (73.3%) +SC-NA had large lymphoid aggregates (LLA) (p = 0.00338). RNA expression was consistent with known acute SARS-CoV2 infection. Hub network analysis showed a tight shift in RNA expression centered around HSPE-1p26, with involvement of known SARS-CoV2 immune mediators like NEAT1. DGE comparative analysis with IBS and acute SARS-CoV2 infection showed higher overlap with acute infection vs. IBS. FGSEA analysis with the same source data demonstrated the same.

**Conclusions:** Our findings establish a syndrome mediated by persistent viral infection (SARS-CoV2 Persistent Intestinal Epithelial Syndrome (SPIES)). We hypothesize that persistent sparse infection drives ongoing immune signaling altering movement and function, creating epithelial and movement effects overlapping with DGBI and IBD

## Introduction

As the impact of acute SARS-CoV-2 infection wanes with prior immune exposure, the cluster of syndromes collectively described as “Long COVID” (LC) remains an ongoing issue and one which has created a substantial burden of disease. The impact of gastrointestinal LC has been substantial, and to date is relatively poorly understood. Prior work has identified a trend towards increasing pediatric inflammatory bowel disease (IBD) since the emergence of COVID^1^, the ability of SARS-CoV-2 to persist in the gut^2, 3^, as well as increased risk of liver disease, demonstrating capacity of this infection to induce durable gastrointestinal complications. Overall, 10-25% of patients report GI symptoms (nausea, anorexia, weight loss, diarrhea, hematochezia) 6 months following infection^4^. Given previous work suggesting rates of persistent viral presence and an overlapping clinical picture^3, 5^, it is possible that a significant fraction of these patients with post-infectious SARS-CoV-2 gastrointestinal symptoms are in fact suffering from pathology driven by persistent viral presence.

Ongoing work into understanding LC has sought to place the condition amongst other conditions marked by relatively minimal histopathological change despite substantial symptom burden, such as irritable bowel syndrome (IBS), myalgic encephalomyelitis/chronic fatigue syndrome (ME/CFS), and other diseases previously referred to as “functional.” Given the broad nature of IBS diagnostic criteria^6^, SARS-CoV-2 infection has been noted as a substantial risk factor for IBS disease formation^7^. While symptoms are broad and varied, overarching work has implicated mechanisms ranging from alteration to serotonin metabolism ^3^, altered coagulation and risk of “microclots” or endothelial injury^8^, or persistent infection, as demonstrated by prior work showing tissue reservoirs of SARS-CoV-2^2, 5^. Involvement of the microbiome has also been implicated, with work demonstrating microbial shifts (decreased butyrate-producing species such as *Faecalibacterium prausnitzii*, increased presence of pathogens such as *Clostridium innocuum* and *Actinomyces naeslundii*)^9^ which correlated with LC symptoms such as fatigue and “brain fog.” In aggregate, there is substantial data suggesting clear pathophysiologic changes associated with GI LC, requiring further understanding of the mechanisms and types of disease manifestations involved.

To further explore underlying drivers of pathology, especially in the context of additional work demonstrating a likely linkage of persistent tissue reservoirs with LC identification of tissue reservoirs in other forms of LC^10^, we have sought to characterize the relevance of persistent SARS-CoV-2 to GI LC symptoms and pathophysiology. Our prior work^2, 5, 11^, initiated after our clinical experience of a proliferation of patients with IBD-like symptoms and “normal” histologic evaluations leading to a presumptive IBS diagnosis, has demonstrated the existence of this phenomenon, which has been confirmed by others^3, 12^. We hypothesized that persistent viral infection was linked to a specific disease phenotype with more discernable endoscopic and histologic changes than typical IBS, and carried out a retrospective clinical study to characterize the symptoms and histology of these patients, coupled with transcriptomics to compare pediatric and young adult patients with persistent SARS-CoV-2 infection to known IBS patients who underwent endoscopy prior to the COVID-19 pandemic.

## Materials & Methods

### Study design and samples

To characterize the clinical and histologic characteristics of persistent intestinal SARS-CoV-2, we reviewed records of patients aged 5-22 years who had undergone clinical, serum, and esophagogastroduodenoscopy for gastrointestinal symptoms at University Hospital Downstate (UHD) in the period June 2020 to June 2023, and who did not possess a histological or laboratory evaluation-supported diagnosis (e.g., IBD, *Helicobacter pylori* infection, celiac disease). Patients with a histologically diagnosable disease or major comorbidity potentially impacting gut health (e.g., severe autism spectrum disorder, cardiac disease) were excluded. Thirty patients were identified, with previously obtained biopsy tissue stained using immunohistochemistry. All patients had duodenal tissue assessed, and in the case of a completed colonoscopy, ileal, cecal, transverse, and rectosigmoid samples were also examined. The following data (if available) was collected for each patient. Age, gender, race/ethnicity, past medical history, symptom profile, history of confirmed or suspected SARS-CoV-2 infection, laboratory evaluation including serum and stool, imaging studies (e.g., magnetic resonance enterography), indication for endoscopy, and vaccination status. Clinical data was compared between positive and negative patients using a t-test for discrete variables and a two-proportion z-test for categorical variables (Prism Graphpad).

### Histopathological profiling

Duodenal, ileal, cecal, and sigmoid colon samples were stained using immunohistochemistry (IHC) for SARS-CoV-2 using a rabbit monoclonal SARS-CoV-2 nucleocapsid antibody (GTX635686, 1:10,000, Histowiz). Stained samples were then provided for independent, blinded pathology review by a single reader to identify positive cases. After positive case identification, the initial clinical samples were provided for blinded pathology review. This review focused on any notable histopathological characteristics for all samples with no knowledge of IHC positivity. Deidentified findings were documented and provided to the remainder of the study team for further analysis. Comparison of histological findings in positive and negative patients was carried out using a two-proportion z-test (Prism Graphpad).

### Transcriptomic sequencing

We selected eight patients with known duodenal immunohistochemical SARS-CoV-2 nucleocapsid antigen positivity for RNA sequencing. To rigorously exclude the possible presence of SARS-CoV-2 in comparison tissue, we identified eight patients from the period 01/01/2015-12/30/2019, i.e., a period preceding the arrival of SARS-CoV-2 in the greater New York City area^13^. To minimize confounding, these patients were selected based on our initial inclusion criteria (absence of histologic or serologic diagnosis), meeting Rome IV criteria for IBS-C or IBS-D^6^, and of matching age, gender, ethnicity, and race to previously identified nucleocapsid antigen-positive patients.

Duodenal tissue was sampled from FFPE-preserved blocks from all study participants. Xylene was used for deparaffinization. RNA was purified using the RNeasy FFPE Kit (Qiagen). Libraries were prepared with the HyperPrep Kit (KAPA) and sequenced on the NextSeq2000 instrument (Illumina) using a P2 flowcell and the PE101 dual-index run format.

### RNA sequencing data analysis

Paired-end reads were trimmed and mapped by STAR ^11^ to the human genome assembly version GRCh38.p14; (BioProject ID: PRJNA31257).. The summed counts of the total samples were converted to counts per million (CPM) in edgeR^14^. Genes with CPM>2 for at least 1/3 of the total sample size were selected. Trimmed mean of M-values (TMM)^15^ was used to estimate normalization factors that account for library size variation between samples. The voom function in limma^16^ was used to estimate the mean-variance relationship of the log-counts to generate a precision weight for each observation and return the voom-normalized feature counts (on the log_2_ scale). Voom-normalized was used for linear modeling with limma.

Weighted gene coexpression network analysis (WGCNA R package)^17^ is a systems biology method used to build gene networks and to detect gene modules and identify genes with high interconnectivity (i.e., hub genes) within modules.

### TF analysis

We conducted transcription factor binding enrichment analyses of all the above DEGs with a P-value <0.05 and for ChEA 2022 and ENCODE TF ChIP-seq 2015 with Enrichr software in R (REF)

#### Comparison of differentially expressed genes (DEG) with other studies

We integrated DEG of this study and 8 independent studies assessing IBS and SARS-CoV-2 epithelial tissue response by calculating Spearman correlation coefficients (rho) of effect sizes (i.e.,log_2_ *FC*) in order to characterize the relationships between them (FIG 1c). Rank-Rank Hypergeometric Overlap analyses^18^ were also performed in the RRHO R package^19^ to identify significant overlap of differential expression lists between pairs of results by determining the degree of statistical enrichment using the hypergeometric distribution. Within each dataset, genes were ranked based on the product of the log_2_ Fold-Change and -log_10_(P-value) from differential expression analyses. The scored gene lists were provided to the RRHO2_initialize function (method “hyper” and log10.ind “TRUE”) and the hypergeometric test −log_10_(P-value) were plotted using the RRHO2_heatmap function (FIG1c).

**Figure 1:**
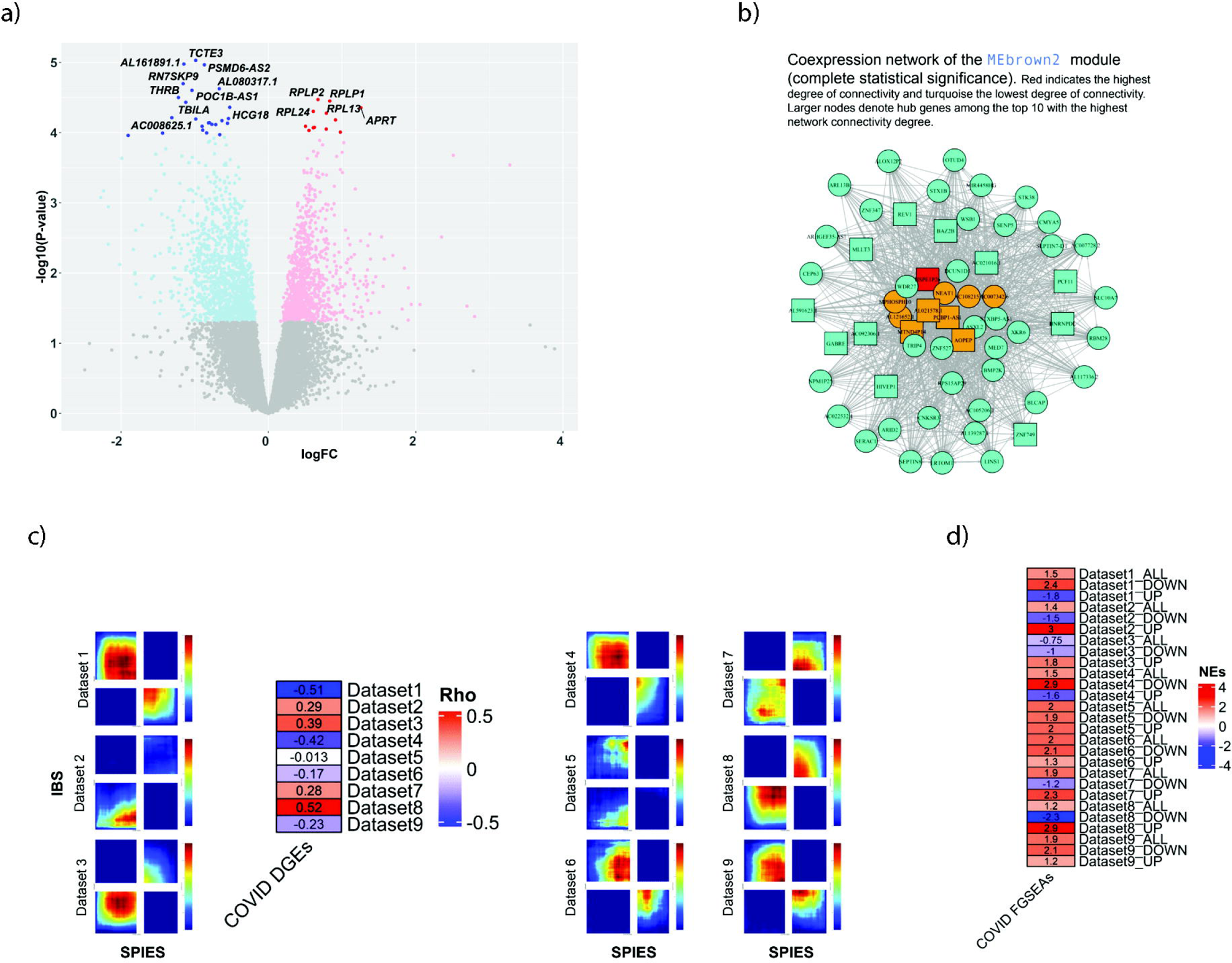
a) Volcano plot representing differential gene expression (DGE) between SC-NA+ and control. b) Hub analysis demonstrating tight linkages and hub gene of HSPE1P26, as well core gene NEAT1, and several others which have been linked with immune response to SARS-CoV2. c) Heatmaps demonstrating comparison between SC-NA+ and IBS patients (left, set 1-3) and various tissues with acute SARS-CoV2 infection (right, set 4-9). d) FGSEAs comparative analysis of up and down regulation between SC-NA+ patients and IBS (1-3)/SARS-CoV2 infection (4-9), demonstrating higher overlap of expression shifts with active infection.

#### Comparison of pathway signatures

The fast gene set enrichment analysis [GSEA] implementation in R [FGSEA]^20^, was used to test concordance of differential expression analysis results with 16,411 gene expression signatures (i.e., gene sets) from curated gene sets available in the Molecular Signatures Database (MSigDB)^21^. For each input dataset, all genes were used for GSEA, and they were ranked based on sign (log_2_ Fold-Change) * -log_10_ (P-value). The gene set size range was set at 15-501. To increase the robustness of results 10,000 permutations were performed. The GSEA returns, for each pathway, the enrichment P-value and the corresponding FDR-adjusted P-value based on a permutation test (i.e., control for the number of the different gene sets tested for enrichment), the enrichment score (ES), the normalized enrichment score (NES; i.e., enrichment score normalized to mean enrichment of random samples of the same pathway size), the nMoreExtreme (i.e., the number of times a random pathway had a more ES), the size (i.e., size of the pathways after removing the not-present genes) and the leading edge (i.e., the leading edge genes for each pathway that drive the enrichment). Pathways with FDR-adjusted P-value < 0.05 were reported.

Normalized enrichment scores from the pathway signatures of this study and nine independent studies were correlated to characterize the relationships between them (FIG1d). Three independent IBS DGE target gene-sets were extracted, as well as 6 independent SARS-CoV-2 acute infection sets from tissue including pulmonary epithelium, airway epithelial cultured cells, cultured hepatocytes, endothelial cells, and iPSC derived alveolar epithelial type 2 cells (**Appendix)**. Overall 27 gene-sets were created (i.e., 3 per dataset) by using: i) the 500 most significant genes (i.e., ALL), ii) the 500 most significant downregulated genes (i.e., log_2_FC<0; DOWN), and iii) the 500 most significant upregulated genes (i.e., log_2_FC>0; UP). These gene-sets were tested for enrichment in DGE of this study (FIG1d).

## Results

The duodenal tissue of 30 patients was assessed via immunohistochemical staining for SARS-CoV-2 nucleocapsid antigen. Patients’ age averaged 14 years old (range 4-20), with a demographic makeup of 56.6% Black, 33.3% Non-Hispanic White, 6.6% Asian, and 3.3% Hispanic White (**Table 1)**. The primary indications for endoscopic evaluation were abdominal pain (90%), nausea/vomiting (63.3%), and weight loss (56.6%), with 20 (66%) having undergone colonoscopic evaluation. Of these patients, 15 (50%) were identified via immunohistochemical (IHC) staining to have persistent SARS-CoV-2 Nucleocapsid antigen, with nine (60%) demonstrating duodenal positivity, two (13.3%) demonstrating terminal ileum positivity, and eight (53.3%) demonstrating colonic positivity. Three patients (20%) showed both duodenal and colonic positivity, and one (6.6%) both duodenal and terminal ileum positivity (**Table 2**). When reviewing only patients who underwent both colonoscopy and esophagogastroduodenoscopy, 11 patients had positive IHC staining, with five (45.5%) having duodenal positivity, two having ileal positivity (18.1%) and eight (72.7%) having colonic positivity (**Table 2**). Three (20%) had received at least a single SARS-CoV-2 vaccine dose in the positive cohort, and eight (53.3%) in the negative cohort (P=0.05). In cases of positive patients with vaccination, it appears that symptoms preceded vaccination. Five (33.3%) of positive patients had a known history of SARS-CoV-2 infection, and none of the negative patients had a documented record of infection (**Table 1**). Of note, data from other health care organizations and validated infection history from the early phase of the COVID-19 pandemic were not available.

**Table 1:**
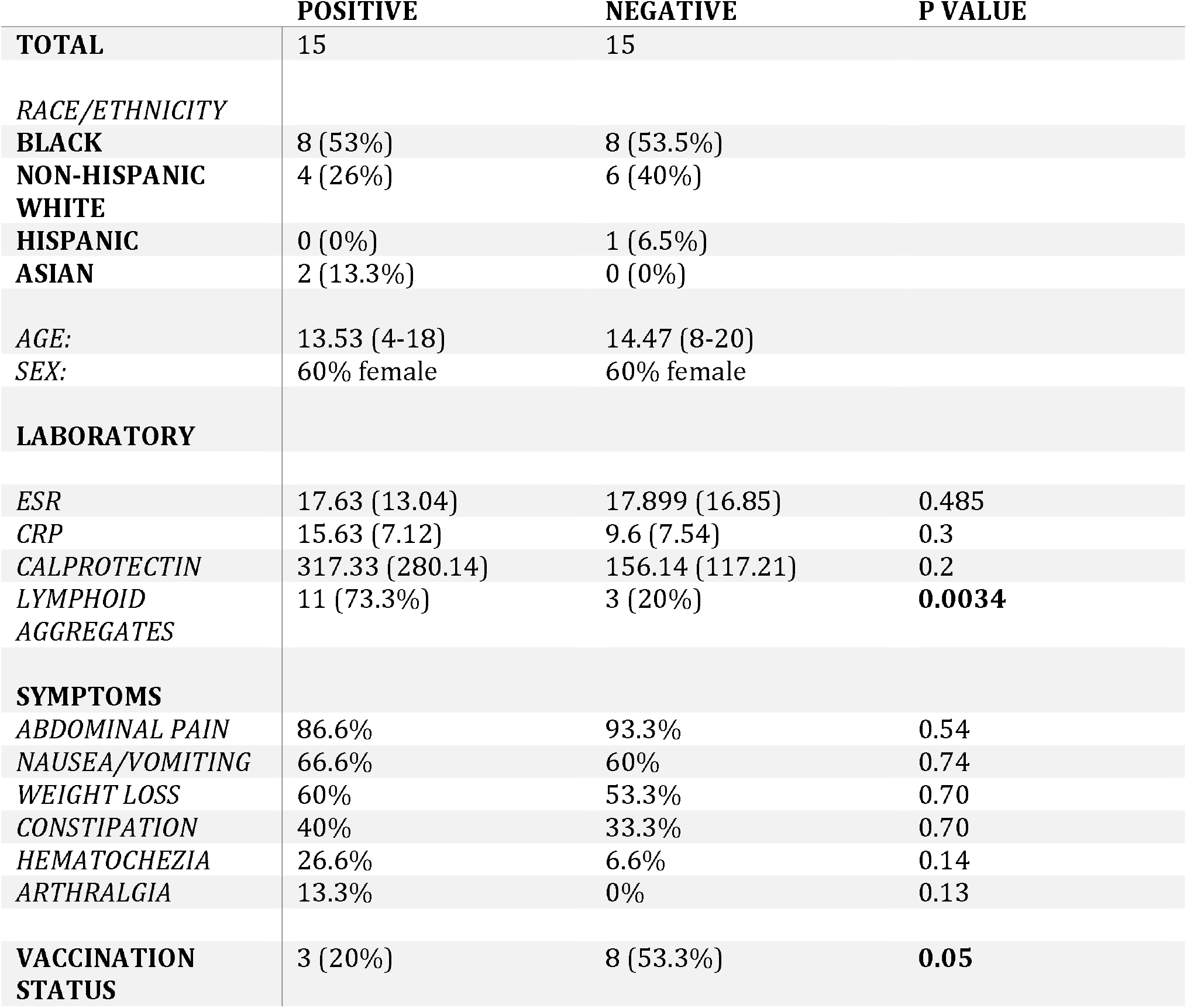
Demographics and characteristics of patients assessed for persistent SARs-CoV-2 Nucleocapsid antigen compared by positivity.

**Table 2:** Clinical Characteristics of Patients with Positive SARS-CoV2 Nucleocapsid Antigen in the Intestine.

|  | DUODENAL | ILEAL | COLONIC |
| --- | --- | --- | --- |
| **TOTAL** | 9 | 2 | 8 |
| *RACE/ETHNICITY* |  |  |  |
| **BLACK** | 6 (66.6%) | 1 (50) | 5 (62.5%) |
| **NON-HISPANIC WHITE** | 3 (33.3%) | 1 (50%) | 1 (12.5%) |
| **HISPANIC** | 0 (0%) | 0 (0%) | 0 (0%) |
| **ASIAN** | 0 (0%) | 0 (0%) | 2 (25%) |
| *AGE:* | 13.53 (4-18) | 16.5 (16-17) | 13.5 (9-18) |
| *SEX:* | 60% female | 100% female | 50% female |
| **LABORATORY** |  |  |  |
| *ESR* | 19 (12.81) | 15 | 17.63 (13.04) |
| *CRP* | 20.45 (7.73) | 14 | 10.31 (7.3) |
| *CALPROTECTIN* | 239.25 (334.12)) | 26.5 | 317.3 (317.8) |
| *LYMPHOID AGGREGATES* | 11 (73.3%) | 2 (100%) | 8 (100%) |
| **SYMPTOMS** |  |  |  |
| *ABDOMINAL PAIN* | 77.8% | 93.3% | 100% |
| *NAUSEA/VOMITING* | 77.8%% | 60% | 50% |
| *WEIGHT LOSS* | 55.5% | 53.3% | 62.5% |
| *CONSTIPATION* | 33.3% | 33.3% | 37.5% |
| *HEMATOCHEZIA* | 22.2% | 6.6% | 37.5% |
| *ARTHRALGIA* | 22.2% | 0% | 0% |
| **COINCIDENT DUODENAL** | N/A | 1 | 3 |
| **COINCIDENT ILEAL** | 1 | N/A | 0 |
| **COINCIDENT COLONIC** | 3 | 0 | 3 |

Patients with positive IHC demonstrated symptoms of abdominal pain (86.6%), nausea/vomiting (66.6%), weight loss (60%), and constipation (40%), although no significant differences were noted between positive and negative patients for these symptoms. 26.6% of positive patients displayed hematochezia compared to only 6.6% of the negative, but this difference was not significant (P=0.14). Similarly, 13.3% of positive patients had arthralgia, and no negative patients reported this, but this difference was not significant (P=0.13). There was no significant variation in symptoms by location of staining positivity, although notably 7/9 (77.7%) patients with duodenal positivity displayed nausea/vomiting compared to 3/6 (50%) patients with a negative duodenal test (P=0.262), and 3/8 (37.5%) of colon IHC-positive patients displayed hematochezia, compared to only 1/7 (14.3%) of non-colon positive patients (P=0.313).

There were no significant differences in serum laboratory evaluation between positive and negative cohorts. One-third of positive patients demonstrated an elevated erythrocyte sedimentation rate [ESR] (mean: 18.1, upper limit normal [ULN]: 10) and 13.3% demonstrated C reactive protein [CRP] elevation (mean: 9.6 mg/L, ULN: 8 mg/L). There was a difference in calprotectin between the positive cohort (mean: 317.3, n= 8 patients) and the negative cohort (mean: 156.4, n=7), although this difference was not significant in our limited cohort (P=0.2). Overall, 20% of positive patients demonstrated elevated fecal calprotectin (**Table 1**).

On histologic review, one patient with duodenal positivity demonstrated signs of mild acute inflammation, and one negative patient demonstrated markers of ileal inflammation suggesting a possible missed diagnosis on the initial pathology review. Notably, patients with positive SARS-CoV-2 IHC staining exhibited the presence of large lymphoid aggregates (LLA) in 11/15 cases (73.3%) with only 3/15 (20%) negative cases displaying such a finding (P=0.00338) (FIG2). When limited solely to patients who underwent esophagogastroduodenoscopy and colonoscopy (n = 20), 100% of positive patients demonstrated LLA in at least one biopsy, compared to 33.3% of negative patients (P=0.0012, PPV: 78.6%, sensitivity: 100%, specificity: 66.6%). LLA distribution was primarily ileocolonic, with only one patient (6.6%) demonstrating duodenal LLA. Of patients with positive duodenal immunohistochemistry and a completed colonoscopy (n=5), 20% had duodenal, 100% had ileal, and 80% had colonic LLA. Of the two patients with positive ileal immunohistochemistry, both had ileal and colonic LLA, but not duodenal. Of patients with positive colonic immunohistochemistry, 12.5% had duodenal LLA, and all had ileal and colonic LLA.

**Figure 2:**
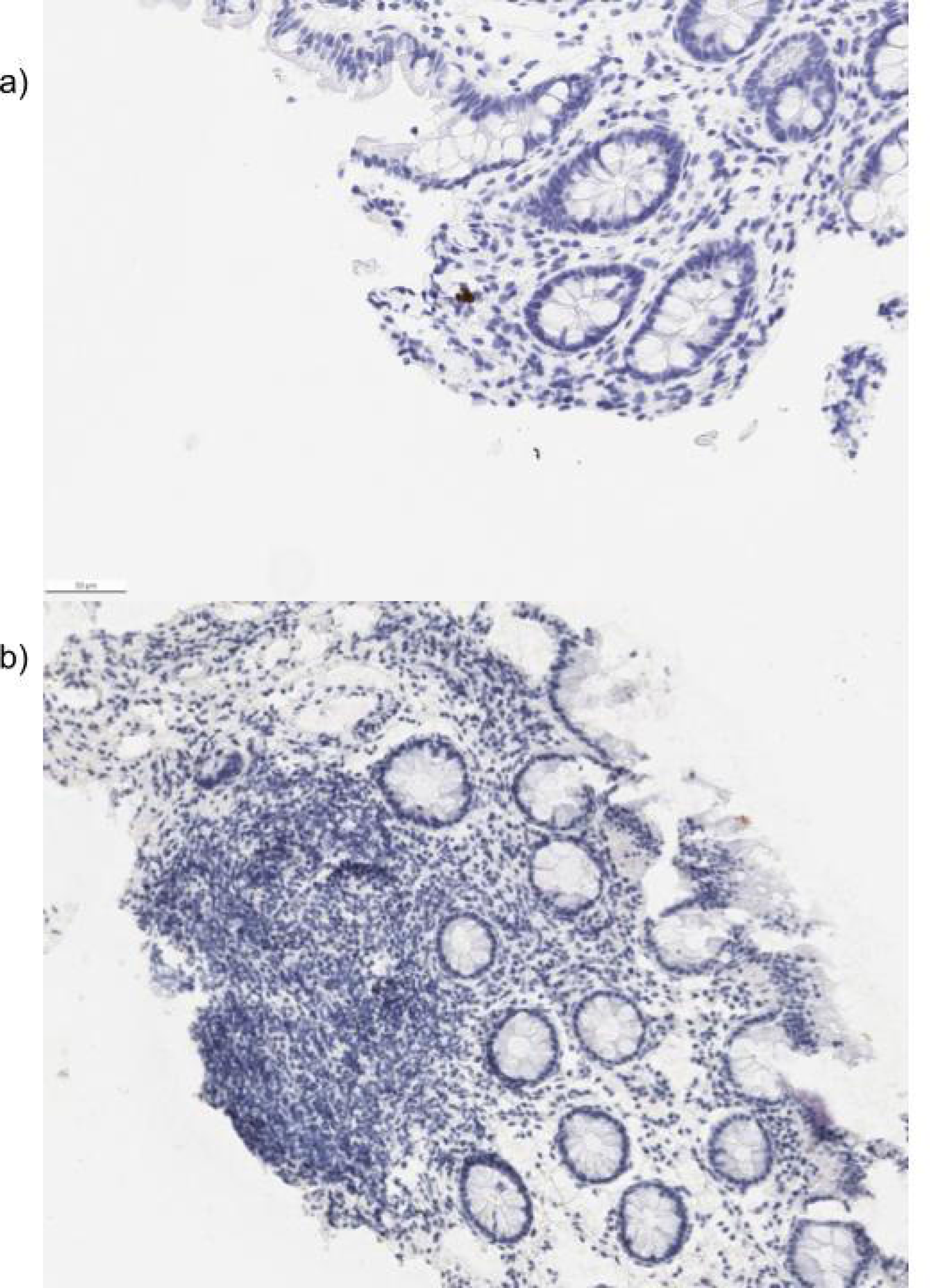
histopathological image of a) +SARS-CoV-2 nucleocapsid staining, and b) germinal lymphoid aggregate associated with presence of + IHC staining.

### RNA Sequencing

The demographic data for patients selected for RNA sequencing is shown in Table 3. Weighted gene correlation network analysis revealed several modules, one of which exhibited complete statistical significance (P<0.00001) and the highest level of intramodular connectivity. Gene HSPE1P2 was the hub gene (**FIG1b**). Comparison with other datasets showed a high level of concordance by the polarity of gene regulation (i.e., upregulated vs. downregulated), but in the case of comparison with IBS data, there more discordance.

**Table 3:** Demographics and characteristics of patients with persistent duodenal SARS-CoV-2 Nucleocapsid Antigen and IBS patients undergoing endoscopy from prior to 2020 for RNA seq comparison.

| **TABLE 3** | **PERSISTENT NA** | **IBS PRE-COVID** |
| --- | --- | --- |
| **TOTAL** | **8** | **8** |
| *RACE/ETHNICITY* |  |  |
| **BLACK** | 5 (62.5%) | 5 (62.5%) |
| **NON-HISPANIC WHITE** | 3 (37.5%) | 3 (37.5%) |
| **HISPANIC** | 0 (0%) | 0 (0%) |
| **ASIAN** | 0 (0%) | 0 (0%) |
| *AGE:* | 15.5 (12-18) | 15.5 (12-18) |
| *SEX:* | 75% female | 75% female |
| **SYMPTOMS** |  |  |
| *ABDOMINAL PAIN* | 87.5% | 87.5% |
| *NAUSEA/VOMITING* | 75% | 25% |
| *WEIGHT LOSS* | 50% | 12.5% |
| *CONSTIPATION* | 37.5% | 12.5% |
| *HEMATOCHEZIA* | 25% | 25% |
| *ARTHRALGIA* | 12.5% | 0% |

The differentially expressed genes (DGE) analysis revealed several genes that are significantly upregulated in the dataset. (**FIG1a)** Notably, the genes RPL24, RPL13, RPLP1, RPLP2, and APRT exhibit positive log fold changes, indicating higher expression levels compared to the control group. These genes are represented as red dots on the volcano plot, demonstrating their statistical significance and increased expression. The identification of these upregulated genes could provide insights into the molecular mechanisms and pathways that are activated in the condition under study. Conversely, the analysis also identified genes that are significantly downregulated, as indicated by negative log fold changes. The genes AL161891.1, TCTE3, PSMD6-AS2, RN7SKP9, AL080317.1, THRB, POC1B-AS1, TBILA, HCG18, and AC008625.1 are marked by blue dots on the volcano plot, highlighting their reduced expression levels. These downregulated genes may play critical roles in the pathways that are suppressed in the condition being investigated. Understanding the patterns of both upregulated and downregulated genes is crucial for comprehending the complex biological processes involved and could aid in identifying potential therapeutic targets or biomarkers.

FGSEA analysis revealed several pathways with significant False Discovery Rate (FDR) values, mostly up-regulated. Importantly, BUSSLINGER DUODENAL_TRANSIT_AMPLIFYING_CELLS” suggests increased activity in gastrointestinal cell proliferation, which aligns with gastrointestinal symptoms observed in many COVID-19 patients **(FIG3)** The “REACTOME EUKARYOTIC TRANSLATION INITIATION” (R-HSA-72649) and “GOCC CYTOSOLIC RIBOSOME” (R-HSA-72689) pathways indicate enhanced protein synthesis, reflecting the cellular response to viral replication, as viruses hijack host machinery for their propagation. The “REACTOME RESPONSE OF EIF2AK4 GCN2 TO AMINO ACID DEFICIENCY” (R-HSA-169732) pathway highlights a stress response mechanism to amino acid depletion, which can occur during intense viral infection as the virus competes for host resources. These enriched pathways illustrate the intricate cellular responses to COVID-19, including increased protein synthesis and stress responses, creating a picture of a persistent response to infection.

**Figure 3:**
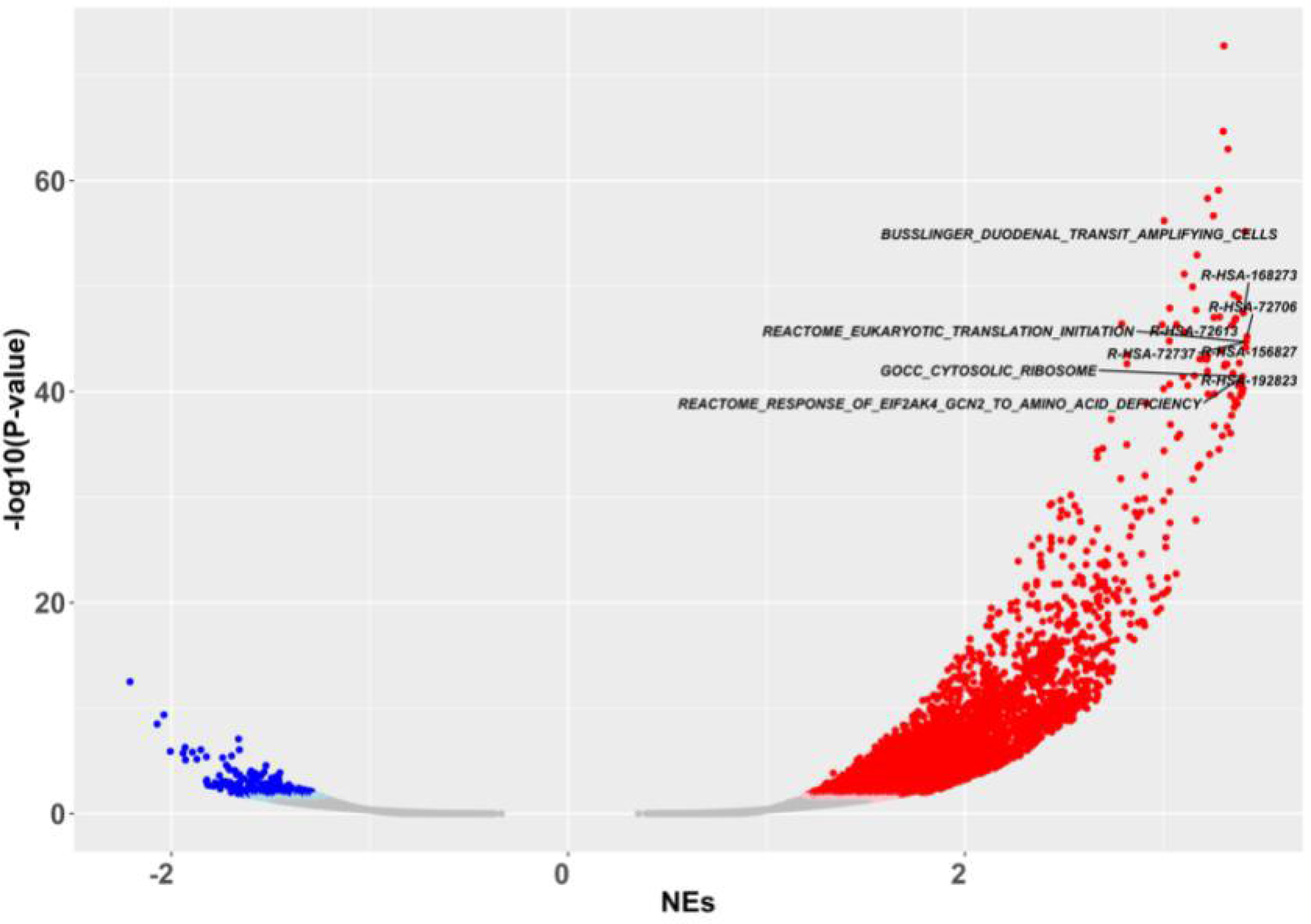
FSGEA analysis of patients with persistent SARS-CoV-2 Nucleocapsid Antigen in comparison with age, gender, and race/ethnically matched IBS patients from prior to 2020.

Weighted Correlation Network Analysis (WGCNA) identified 26 modules (**FIG1b)**. Notably, modules such as MEmagenta, MEbrown4, MEbrown2, and MEdarkolivegreen exhibit high statistical significance and negative log fold changes, while MEsaddlebrown and MEfirebrick4 exhibit positive log fold changes. Finally, the MEbrown2 module (genes=59) had 17 genes that were also nominally significant DEGs (p<0.05), with the hub gene HSPE1P26 also being a nominal DEG. Additionally, the heatmap analysis of differentially enriched transcription factors (TFs) in Fig. xx highlights notable TFs. Specifically, MYC, CTCF, and XRN2 were the most significant, depicted in red. Other notable transcription factors with significant expression changes include SOX2, EGR1, NANOG, ESR1, and FOXA1. The clustering and color coding suggest that these transcription factors may be involved in the regulatory mechanisms and pathways influenced by COVID-19 infection. Understanding these changes is essential for comprehending the transcriptional landscape and potential impacts on cellular function during the infection.

The Spearman correlation coefficient (rho) between the current DEGs and nine independent datasets ranged from -0.51 to 0.52. Notably, Dataset1 (colonic tissue, IBS) and Dataset4 (Lung tissue, SARS-CoV-2 infection), showed strong negative correlations (−0.51 and -0.42, respectively), indicating a divergent gene expression profile compared to the other datasets. In contrast, Dataset8 (human umbilical vein endothelial cells, SARS-CoV-2 infection) showed the highest positive correlation (0.52), suggesting a similar gene expression pattern with other datasets. These correlations highlight the variability and potential commonalities in gene expression responses to COVID-19 across different studies. Moreover, Rank-Rank Hypergeometric Overlap (RRHO) analyses along with rho between FGSEA results of this study and the nine independent ones revealed similar patterns (**FIG1c)**.

Lastly, the 27 curated pathways, as described in the methods, were tested for enrichment in the DEG of this study. Notably, the normalized enrichment scores (NES), ranging from -4 to 4, are color-coded from blue (negative NES) to red (positive NES). Datasets such as Dataset1_ALL (1.5), Dataset3_UP (3), and Dataset8_UP (2.3) show strong positive enrichment, indicating significant pathway activation. Conversely, Dataset3_DOWN (−3), Dataset2_DOWN (−1.5), and Dataset4_DOWN (−1.6) exhibit negative NES values, reflecting pathway suppression. These enrichment patterns highlight the varied molecular responses to COVID-19 across different datasets, emphasizing the differential regulation of pathways involved in the disease (**FIG1d)**.

## Discussion

In the manuscript we present clear clinical, histologic, and RNA transcriptomic evidence of a novel SARS-CoV-2 syndrome predicated on a local inflammatory response to persistent viral presence in the intestinal epithelium. We have titled these phenomena “SARS-CoV-2 Persistent Intestinal Epithelial Syndrome (SPIES).” This novel disease type presents with a clinical picture frequently suggestive of mild-moderate Crohn’s Disease, with symptoms of abdominal pain and weight loss being common. Other symptoms appear to have some linkage with the location of persistent infection, with duodenal disease generating more nausea and vomiting, and colonic disease being linked with a risk of hematochezia, although further data is needed to identify if this early association continues. Our results demonstrate that this condition is common in our patient population (50% of all patients undergoing endoscopy who did not have an endoscopic or histologic diagnosis made). Given that prior work has estimated diagnostic yield of endoscopy in children as between 40-50%^22, 23^, this suggests that up to a quarter of all pediatric and young adult patients undergoing endoscopic evaluation subsequent to the emergence of SARS-CoV-2 may in fact have symptoms induced by SPIES.

We define this condition as follows:

- Gastrointestinal symptoms including abdominal pain, weight loss, nausea, and hematochezia, of greater than 2 months duration.
- endoscopic evaluation demonstrating notably enlarged lymphoid aggregates and presence of persistent SARS-CoV-2 on IHC.

While we so far have demonstrated this phenomenon in pediatric and young adult patients, literature from adults demonstrates that persistent viral presence remains a phenomenon associated with post viral symptoms^3^. Based on our sequencing comparison, this phenomenon is distinct from IBS (and likely other diseases of gut brain interaction) although patients with SPIES may currently be felt to have IBS due to the absence of other diagnosable conditions. It is unclear from our initial cohort the expected duration of this condition, but we have observed that almost all patients over time demonstrated some improvement in symptoms. In our patient cohort, the bulk of patients were managed symptomatically, with a mix of laxatives, low dose SSRI, mirtazapine, and 5-ASA therapies providing mild to moderate improvement, although diversity of therapeutic approach did not allow for any statistical comparisons in terms of treatment efficacy. While no significant differences in symptoms were noted between cohorts, this is confounded by small sample size, and a probability of missed persistent SARS-CoV-2 given the sparse nature of viral presence.

In aggregate, both our histologic and RNA transcriptional data align in demonstrating a condition marked by low-grade, persistent inflammation due to the retained presence of SARS-CoV-2 virus and a consistent immune response against it. SARS-CoV-2 has known gut tropism, entering the intestinal epithelium via ACE-2 receptors^24^. Presence of tissue reserves in the gut have also been established by prior work^2, 25^. Our finding of a linked histological marker of large lymphoid aggregates is consistent with a local immune response to ongoing viral presence^26^, and the significant association with the presence of IHC confirmed SARS-CoV-2 is a useful mechanism for identifying patients for whom IHC has utility, much like similar endoscopic and histologic findings in determining whether *H. pylori* staining is indicated^27^. Lymphoid aggregates on their own are a nonspecific finding consisting of clusters of B, T, and supporting immune cells. They have been linked with inflammatory response, and allergy, and have previously been tied to severity of gastrointestinal symptoms^28-30^. In this case, it seems highly likely that persistent viral presence is driving lymphoid expansion in the gut.

Analysis of our sequencing results demonstrates an unusually coherent network analysis (WCGNA^17^), as well as a unique pattern of differentially expressed genes and pathways in comparison to patients with IBS who underwent endoscopy prior to 2020, ensuring no plausible presence of SARS-CoV-2 in the epithelium (**FIG1a, FIG1b**). The network hub gene, *HSPE1P2*, has an upregulation known to be heavily linked with T cell activation, a central component of cellular SARS-CoV-2 response^31, 32^. In association with the histologic evidence of lymphoid aggregates, this result aligns well with observed clinical data. T cell activation has also been linked with other forms of long COVID^33^. The remainder of the WCGNA analysis also aligns with both the presence of SARS-CoV-2 and an inflammatory response, with *NEAT1* upregulation also noted in SARS-CoV-2 infection and linked with cytokine increase^34^. *NEAT1* expression has also previously been associated with upregulation of IL-4, IL-5, IL-13, IL-17, and increases level of INF-γ in CD4^+^ cells.

SARS-CoV-2 has been theorized to drive IBS presentation as a secondary effect, however, we hypothesize that persistent intestinal epithelial presence differs substantially from IBS, and that a subset of patients receiving IBS diagnoses after SARs-CoV-2 are in fact SPIES patients. In our comparison of SPIES patients with previously published IBS transcriptomics data and sequencing derived from tissue with acute SARS-CoV-2 infection, we note a closer fit with SARS-CoV-2 infection based on differential gene expression, and a substantially higher alignment with acute SARS-CoV-2 infection when deploying a gene set enrichment analysis (**FIG1d**). Our findings suggest that SPIES is a mosaic of an IBS gene regulatory signature dominated by host response to persistent SARS-CoV-2 infection. These findings highly suggest that the primary driving factor of symptoms in this condition is immune response to viral presence, although there are some overlapping programmatic signatures with IBS.

SARS-CoV-2 has demonstrated a capacity to inhibit local immune response to a degree that facilitates viral replication^30^. Our transcriptomics results reveal several altered pathways which may support ongoing immune constraints allowing for persistent infection, most specifically a downregulation of THRB (thyroid receptor hormone β) which can constrain interferon-γ and other inflammatory modulators^35^. We theorize that persistent infection is sustained in individuals with altered local antiviral response for any reason (including SARS-CoV-2 modulatory effects), allowing SARS-CoV-2 to maintain ongoing -if sparse-infections with localized mild immune signaling resulting in an inflammatory, pro-proliferative state which impairs intestinal function and increases pain sensitivity. This inflammatory state may alter absorptive capacity, as well as generating symptoms limiting dietary intake, resulting in presenting symptoms often consistent with Crohn’s Disease. It appears from our data that SARS-CoV-2 vaccination may be protective in this condition, although a larger epidemiological study design will be better suited to address this.

Our study has several limitations. The nature of FFPE samples and the retrospective examination constrain the assessment of the natural history of the disease. Duodenal acute SARS-CoV-2 infected tissue sequencing sources were not able to be found, likely due to low use of esophagogastroduodenoscopy during acute SARS-CoV-2-infection. However, using multiple cell types allows for the identification of common features, and our alignment with gene expression in varied cell types with acute infection strongly supports an ongoing persistent infectious process. Recruitment of large participant populations is challenging in pediatric studies.

Our findings demonstrate the appearance of a novel infection phenotype and a related clinical syndrome. The condition is marked by specific intestinal epithelial changes, notably large lymphoid aggregates, and appears to be mediated by a low-intensity localized inflammatory response in the intestine. It remains unclear if this phenomenon is unique to SARS-CoV-2, and as of this time, additional investigation into the pathophysiology and treatment approaches is indicated.

## Acknowledgments

Erica Licari, Po-Ting Lu, Maria Aguilera, Arthi Ramu, Jenny Libien, David Christini.

## Data availability

on GEO pending acceptance

